# Association of SARS-CoV-2 Genomic Load with COVID-19 Patient Outcomes

**DOI:** 10.1101/2020.07.02.20145151

**Authors:** Ioannis M. Zacharioudakis, Prithiv J. Prasad, Fainareti N. Zervou, Atreyee Basu, Kenneth Inglima, Scott A. Weisenberg, Maria E. Aguero-Rosenfeld

**Affiliations:** Division of Infectious Diseases and Immunology, Department of Medicine, NYU Grossman School of Medicine, New York, NY, USA; Department of Pathology, NYU Grossman School of Medicine, New York, NY, USA

**Keywords:** SARS-CoV-2, COVID-19, RT-PCR, Viral Load

## Abstract

**Rationale:** The Infectious Diseases Society of America has identified the use of SARS-CoV-2 genomic load for prognostication purposes as a key research question.

**Objectives:** We explored the SARS-CoV-2 genomic load as a risk factor for adverse patient outcomes.

**Methods:** A retrospective cohort study among adult patients admitted to the hospital between March 31^st^ to April 10^th^, 2020 with COVID-19 pneumonia was conducted. We segregated patients into 3 genomic load groups: low (Cycle threshold (Ct) ≥35), intermediate (25<Ct<35), and high (Ct≤ 25) using real-time polymerase chain reaction.

**Measurements:** A composite outcome of death, intubation, and/or extracorporeal membrane oxygenation was used. Secondary outcomes included the severity of pneumonia on admission, as measured by the Pneumonia Severity Index (PSI).

**Main Results:** Of 457 patients with COVID-19 pneumonia from March 31^st^ to April 10^th^, 2020, 316 met inclusion criteria. Included patients were followed for a median of 25days (IQR 21-28). High genomic load at presentation was associated with higher Charlson Comorbidity Index (p=0.005), transplant recipient status (p<0.001), and duration of illness less than 7 days (p=0.005). Importantly, patients with high genomic load were more likely to reach the primary endpoint (p=0.001), and had higher PSI scores on admission (p=0.03). In multivariate analysis, a high genomic load remained an independent predictor of the primary outcome. Results remained significant in sensitivity analyses.

**Conclusions:** Our findings suggest that a high genomic load of SARS-CoV-2 at the time of admission is an independent predictor of adverse outcomes, that above and beyond age, comorbidity, and severity of illness on presentation, may be used to risk-stratify patients, and call for a quantitative diagnostic assay to become available.

## Introduction

The initial outbreak of Coronavirus Disease 2019 (COVID-19) caused by the SARS-CoV-2 virus in Wuhan City, Hubei Province, China was reported to the World Health Organization (WHO) on December 31^st^, 2019 (1). Since its recognition, there have been 2.5 million cases of COVID-19 reported worldwide (2). New York city has become the epicenter of disease with more than 130,000 confirmed cases and 9,000 deaths, as of April 20^th^, 2020 (3). The surge in infections worldwide has overwhelmed health care systems, and identifying patients likely to need and benefit from intensive care is critically important (4). Host factors, including age and certain comorbidities, have been associated with mortality (5–9). Viral factors might also influence disease severity. However, their role in COVID-19 outcomes is to date unclear (10).

We aimed to study the association of SARS-CoV-2 genomic load in nasopharyngeal samples with clinical outcomes. We used the Cycle threshold (Ct) value, the number of amplification cycles needed to yield a positive fluorescent signal in a real-time reverse transcription-polymerase chain reaction test (RT-PCR), as a surrogate for viral load.

## Methods

### Study Design, Setting and Participants

We conducted a retrospective cohort study at the NYU Langone Medical Center, a tertiary academic medical center in New York City. We evaluated a convenience sample of patients who presented to the emergency department between March 31^st^ and April 10^th^ of 2020 and tested positive for SARS-CoV-2. Hospitalized adult patients were included in the analysis if they demonstrated clinical findings of pneumonia defined as a subjective or documented fever on admission >100.4° Fahrenheit, shortness of breath or hypoxia (blood oxygen saturation <90%), or new or worsening cough from baseline as well as radiographic findings of viral pneumonia. New or worsening airspace or interstitial opacities on chest X-Ray or Computed Tomography compared to baseline imaging were considered suggestive of viral pneumonia. We excluded patients who were tested >24 hours into the admission, as our goal was to study the association of the genomic load at the time of admission to the hospital, with patient outcomes.

### Real-time reverse transcriptase Polymerase Chain Reaction (RT-PCR) Assay

On March 31^st^, 2020, our microbiology laboratory adopted the Cepheid Xpert^®^ Xpress SARS-CoV-2 assay for in-house diagnosis of COVID-19. This is a rapid RT-PCR test intended for the qualitative detection of ribonucleic acid (RNA) from SARS-CoV-2 in individuals suspected of COVID-19. After processing, the specimens are run on the GeneXpert^®^ platform, which automates and integrates sample preparation, nucleic acid extraction and amplification, and detection of the target sequences. The Xpert^®^ Xpress SARS-CoV-2 test is only for use under the Food and Drug Administration’s Emergency Use Authorization as a qualitative result (11). The assay detects two nucleic acid targets, namely N2 and E, and reports the Ct values. The Ct values provide a semiquantitative measure of genomic load, with an inverse relationship between genomic load and Ct value (12). The N2 target is specific for SARS-CoV-2, while the E nucleic acid can also be found in SARS-CoV-1. A positive assay result implies that either N2 and E or N2 target alone were detected, whereas detection of the E nucleic acid alone is considered a presumptive positive result (the latter were excluded from this study). A sample is considered negative by the instrument if it has a Ct value beyond the acceptable valid range and endpoint above the minimum setting for the N2 and E targets, with a positive sample processing control. The lowest limit of detection for this assay is 250 copies/mL.

### Outcome Measures and Follow-up

The primary study outcome was the association of the genomic load in patients admitted to the hospital with COVID-19 pneumonia with disease outcomes. We used a composite outcome of death or discharge to hospice care, use of mechanical ventilation or extracorporeal membrane oxygenation (ECMO). The composite outcome was selected to acknowledge that a minority of patients who receive mechanical ventilation for COVID-19 would be extubated successfully (5, 13, 14). In a sensitivity analysis, a modified composite outcome that also included acute respiratory distress syndrome (ARDS) was studied. We defined ARDS as acute-onset hypoxemia (partial pressure of arterial oxygen to the fraction of inspired oxygen [Pao2: Fio2], <300) with bilateral pulmonary opacities on chest imaging, not explained by volume overload, using the Berlin criteria (15). The Berlin definition requires that arterial blood gases are performed under a positive end-expiratory pressure (PEEP) ≥5cm H_2_O. There is a debate on whether patients that receive oxygenation via a High-Flow Nasal Cannula (HFNC) meet this definition (16). Studies have demonstrated that HFNC is associated with significant positive airway pressure in volunteers, exceeding the threshold of 5cm in those receiving HFNC at a rate of 40L or more. As such, we considered those patients eligible if they otherwise met the Berlin criteria. Outcomes could be immediate, or at any time in the hospital course. All patients were followed up until April 30^th^, 2020, at which point patient data were censored.

### Data Collection

We reviewed electronic medical records of included patients and extracted the following information: demographic characteristics, Body Mass Index (BMI), defined as the patient’s weight in kilograms divided by the square of height in meters, smoking history and respiratory comorbidities including cystic fibrosis, bronchiectasis, interstitial lung disease, chronic obstructive pulmonary disease (COPD), asthma. History of human immunodeficiency virus (HIV), solid organ (SOT) or hemopoietic stem cell transplant (HSCT), hematologic malignancies, chronic systemic steroid or use of other immunosuppressive or immunomodulatory agents were also extracted. The Charlson Comorbidity Index (CCI) was calculated to assess for comorbid conditions. The Pneumonia Severity Index (PSI) was also calculated. The PSI is a validated prediction tool of outcomes in patients with community-acquired pneumonia that uses clinical, laboratory, and radiographic findings. Patients are divided into 5 classes with a higher class at the time of admission being associated with worse outcomes (17). Duration of symptoms prior to presentation, the C-Reactive Protein (CRP), and radiographic findings were also extrapolated.

### Statistical Analysis

We arbitrarily categorized Ct values to classify patients into 3 SARS-CoV-2 genomic load status groups: low, ≥35, intermediate, 25–35, and high, ≤25. A difference of 3.0 in Ct values represents, in general, an approximate order of magnitude difference in genomic load. We compared the patients in the 3 genomic load groups based on demographic characteristics, BMI, smoking history, Charlson Comorbidity Index, pulmonary comorbidities, immunosuppressive diseases, duration of symptoms, Pneumonia Severity Index, CRP and fever at the time of admission using the Chi-square test. A multivariate logistic regression analysis was performed to examine the association of SARS-CoV-2 genomic load with the primary composite outcome adjusted for patient demographics, BMI, smoking history, pulmonary comorbidities, transplant status, Pneumonia Severity Index, and duration of symptoms. The marginal method was used to estimate the probability of the composite outcome among patients with low, intermediate, and high genomic loads when all the other variables were fixed at their means (18). We performed sensitivity analyses adding ARDS to the composite outcome (19), and evaluating the primary outcome using the tertiles of the Ct values observed as breakpoints for classifying the genomic load. All calculations were performed using the Stata v14.2 software package (Stata Corporation, College Station, TX). Results are presented as proportions with 95% confidence intervals (CIs), medians with interquartile ranges (IQR), and odds ratios (ORs) with 95% CIs. A *P* value of <0.05 was considered statistically significant. This study was approved with a waiver of informed consent by the New York University Institutional Review Board.

## Results

457 patients who presented to our emergency department from March 31^st^ to April 10^th^, 2020, tested positive for SARS-CoV-2. Of those admitted 314 met the inclusion criteria and were included in the final analysis. The Flow Chart is presented in Figure 1. Among the included patients, the median age was 64 years (IQR 54-72), 205 (65.3%) were male, 140 (44.6%) were white, with a median BMI of 28.3 (IQR 25.1-32.3). In terms of comorbidities, median Charlson Comorbidity Index was 3 (IQR 1-5) and 117 patients (37.3%) were obese, as defined with a BMI of equal or more than 30kg/m^2^. Additionally, 50 patients (15.9%) had at least one pulmonary comorbidity, 72 (23.5%) were active or former smokers, 21 (6.7%) were transplant recipients (20 SOT, 1 HSCT) and 4 had HIV (3 of them virologically suppressed). Baseline patient characteristics are presented in Table 1. The median duration of symptoms prior to presentation was 7 days (IQR 5-10). Chest imaging demonstrated multifocal opacities in 242 patients (77.1%). Using the Pneumonia Severity Index, 9 patients were classified in class I (2.9%), 78 patients in II (24.8%), 84 patients in III (26.8%), 102 patients in IV (32.5%) and 41 in V (13%).

**Table 1.** Patient characteristics.

| **Demographics/Patient Characteristics** | **No. of patients (%) or Median (IQR)** |
| --- | --- |
| **Age** | 64 (54-72) |
| **Gender** |  |
| Female | 109 (34.7%) |
| Male | 205 (65.3%) |
| **Race** |  |
| White | 140 (44.6%) |
| Afr. American/Black | 41 (13.1%) |
| Asian | 35 (11.2%) |
| Hispanic | 16 (5.1%) |
| Other/Unknown | 82 (26.0%) |
| **BMI** | 28.3 (25.1-32.3) |
| **Obesity (BMI ≥30)** | 117 (37%) |
| **Pulmonary Comorbidities** | **No. of patients (%)** |
| Asthma | 23 (7.3%) |
| COPD | 12 (3.8%) |
| OSA | 22 (7.0%) |
| Any pulmonary comorbidity | 50 (15.9%) |
| **Smoking (current/ former)** | 72 (23.5%) |
| **Transplant** | 21 (6.7%) |
| **Charlson Comorbidity Index** | 3 (1-5) |
| **Nursing Home Residents** | 16 (5.1) |

<sup>1</sup>. **Footnote:** BMI: Body Mass Index, IQR: Interquartile range, No: number.

**Figure 1.**
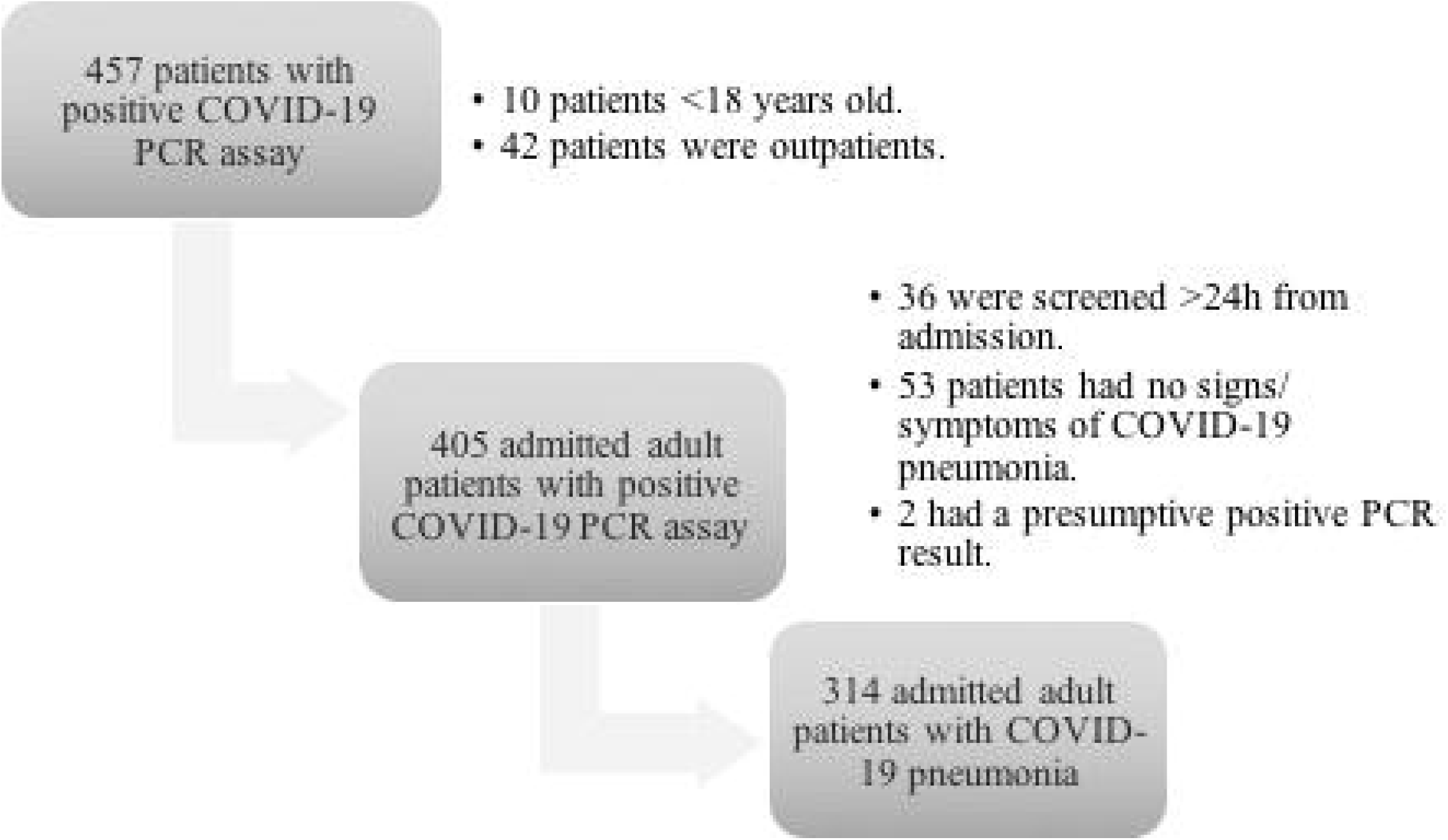
Flow Chart.

Of the 314 included patients, 96 (30.6%) were categorized into the low, 157 (50.0%) into the intermediate, and 61 (19.4%) into the high SARS-CoV-2 genomic load category. Demographic characteristics and comorbidities across the genomic load groups are presented in Table 2. Patients with high genomic loads had higher Charlson Comorbidity scores (p=0.005), were more likely to be transplant recipients (p<0.001), and had a significantly shorter duration of symptoms (p=0.005). There was a trend towards higher genomic load among patients older than 65 (p=0.09). There was no statistically significant difference in CRP values (p=0.43) or presence of fever (>100.5°F) on admission (p=0.28) in patients with high versus low genomic load. The Pneumonia Severity Index was significantly higher in patients with high genomic loads (p=0.03). All patients were followed until April 30^th^, 2020, or until a primary endpoint was reached. The follow-up period was a median of 25 days (IQR 21-28). At the end of follow up, the composite outcome of death/hospice, intubation, or ECMO was reached by 74 patients (23.6%). Median time to primary outcome was 3.5 days (IQR 1-6). On the day of censoring, 309 patients (98.4%) had either reached the primary outcome or were discharged. Compared with patients with low genomic load, patients with high genomic load had a significantly higher unadjusted risk to reach the composite outcome of death, intubation, or ECMO (p=0.001). Table 3 summarizes clinical outcomes, by SARS-CoV-2 genomic load category.

**Table 2.**
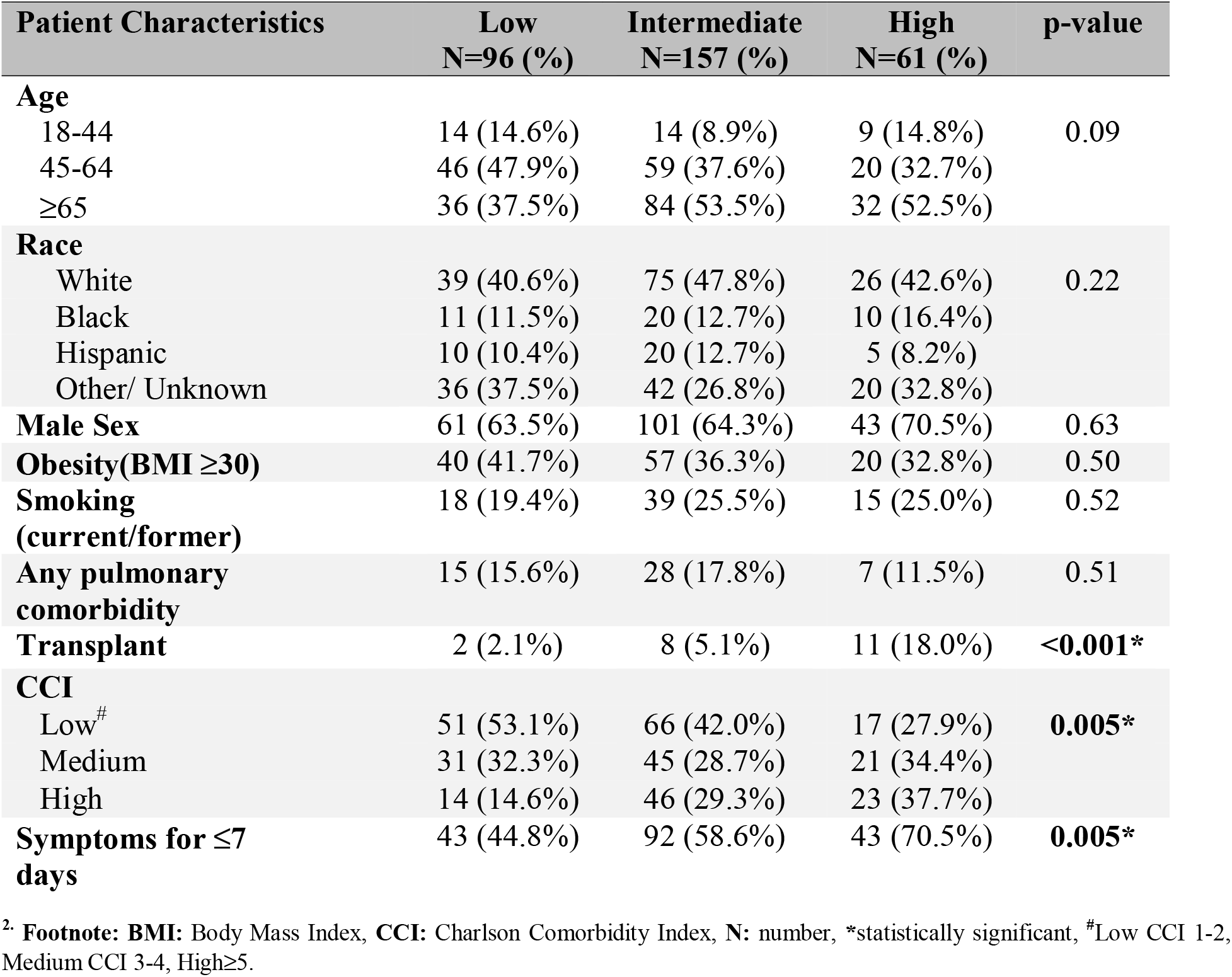
Association of SARS-CoV-2 Genomic Load with Patient Characteristics.

| **Patient Characteristics** | **Low**<br>**N=96 (%)** | **Intermediate**<br>**N=157 (%)** | **High**<br>**N=61 (%)** | **p-value** |
| --- | --- | --- | --- | --- |
| **Age** |  |  |  |  |
| 18-44 | 14 (14.6%) | 14 (8.9%) | 9 (14.8%) | 0.09 |
| 45-64 | 46 (47.9%) | 59 (37.6%) | 20 (32.7%) |  |
| ≥65 | 36 (37.5%) | 84 (53.5%) | 32 (52.5%) |  |
| **Race** |  |  |  |  |
| White | 39 (40.6%) | 75 (47.8%) | 26 (42.6%) | 0.22 |
| Black | 11 (11.5%) | 20 (12.7%) | 10 (16.4%) |  |
| Hispanic | 10 (10.4%) | 20 (12.7%) | 5 (8.2%) |  |
| Other/ Unknown | 36 (37.5%) | 42 (26.8%) | 20 (32.8%) |  |
| **Male Sex** | 61 (63.5%) | 101 (64.3%) | 43 (70.5%) | 0.63 |
| **Obesity(BMI ≥30)** | 40 (41.7%) | 57 (36.3%) | 20 (32.8%) | 0.50 |
| **Smoking**<br>**(current/former)** | 18 (19.4%) | 39 (25.5%) | 15 (25.0%) | 0.52 |
| **Any pulmonary**<br>**comorbidity** | 15 (15.6%) | 28 (17.8%) | 7 (11.5%) | 0.51 |
| **Transplant** | 2 (2.1%) | 8 (5.1%) | 11 (18.0%) | <0.001* |
| **CCI** |  |  |  |  |
| Low<sup>#</sup> | 51 (53.1%) | 66 (42.0%) | 17 (27.9%) | **0.005\*** |
| Medium | 31 (32.3%) | 45 (28.7%) | 21 (34.4%) |  |
| High | 14 (14.6%) | 46 (29.3%) | 23 (37.7%) |  |
| **Symptoms for ≤7**<br>**days** | 43 (44.8%) | 92 (58.6%) | 43 (70.5%) | **0.005\*** |

<sup>2</sup> **Footnote:** BMI: Body Mass Index, CCI: Charlson Comorbidity Index, N: number, \*statistically significant, <sup>#</sup>Low CCI 1-2, Medium CCI 3-4, High≥5.

**Table 3.**
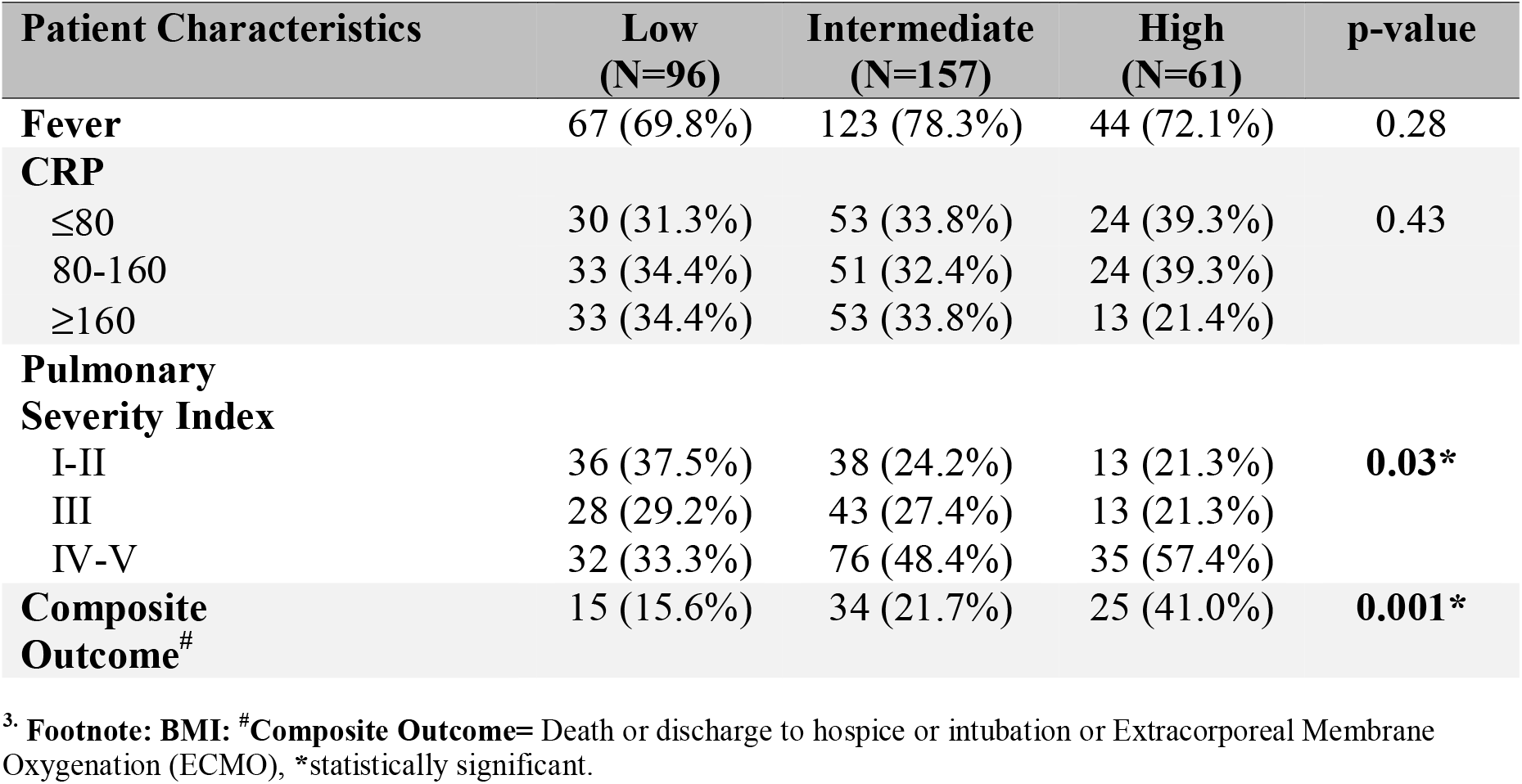
Association of SARS-CoV-2 Genomic Load with Patient Outcomes.

| **Patient Characteristics** | **Low**<br>**(N=96)** | **Intermediate**<br>**(N=157)** | **High**<br>**(N=61)** | **p-value** |
| --- | --- | --- | --- | --- |
| **Fever** | 67 (69.8%) | 123 (78.3%) | 44 (72.1%) | 0.28 |
| **CRP** |  |  |  |  |
| ≤80 | 30 (31.3%) | 53 (33.8%) | 24 (39.3%) | 0.43 |
| 80-160 | 33 (34.4%) | 51 (32.4%) | 24 (39.3%) |  |
| ≥160 | 33 (34.4%) | 53 (33.8%) | 13 (21.4%) |  |
| **Pulmonary**<br>**Severity Index** |  |  |  |  |
| I-II | 36 (37.5%) | 38 (24.2%) | 13 (21.3%) | **0.03\*** |
| III | 28 (29.2%) | 43 (27.4%) | 13 (21.3%) |  |
| IV-V | 32 (33.3%) | 76 (48.4%) | 35 (57.4%) |  |
| **Composite**<br>**Outcome<sup>#</sup>** | 15 (15.6%) | 34 (21.7%) | 25 (41.0%) | **0.001\*** |

<sup>3</sup>. Footnote: BMI: <sup>#</sup>Composite Outcome= Death or discharge to hospice or intubation or Extracorporeal Membrane Oxygenation (ECMO), \*statistically significant.

In the multivariate model, controlling for patient demographics, BMI, pulmonary comorbidities, smoking and transplant history, duration of symptoms, and Pneumonia Severity Index, high genomic load remained an independent risk factor for the composite outcome (OR 2.89; 95%CI 1.7-14.5); p= 0.004). The margins analysis indicated that the average probability of the composite outcome would be 32% (95% CIs 0.16-0.48) if everyone had a high genomic load compared to 9% (95% CIs 0.02-0.15) if everyone had a low genomic load (Figure 2). Among patients with a high Pneumonia Severity Index, the expected probability of the composite outcome was 55% (95% CIs 0.38-0.73) for those with a high genomic load as opposed to 20% (95% CIs 0.06-0.34) for those with a low genomic load (Figure 3). In sensitivity analyses, the association of high genomic load with adverse outcomes remained significant when ARDS was included in the composite outcome (p=0.05) and when varying Ct classification breakpoints were used (p=0.004).

**Figure 2.**
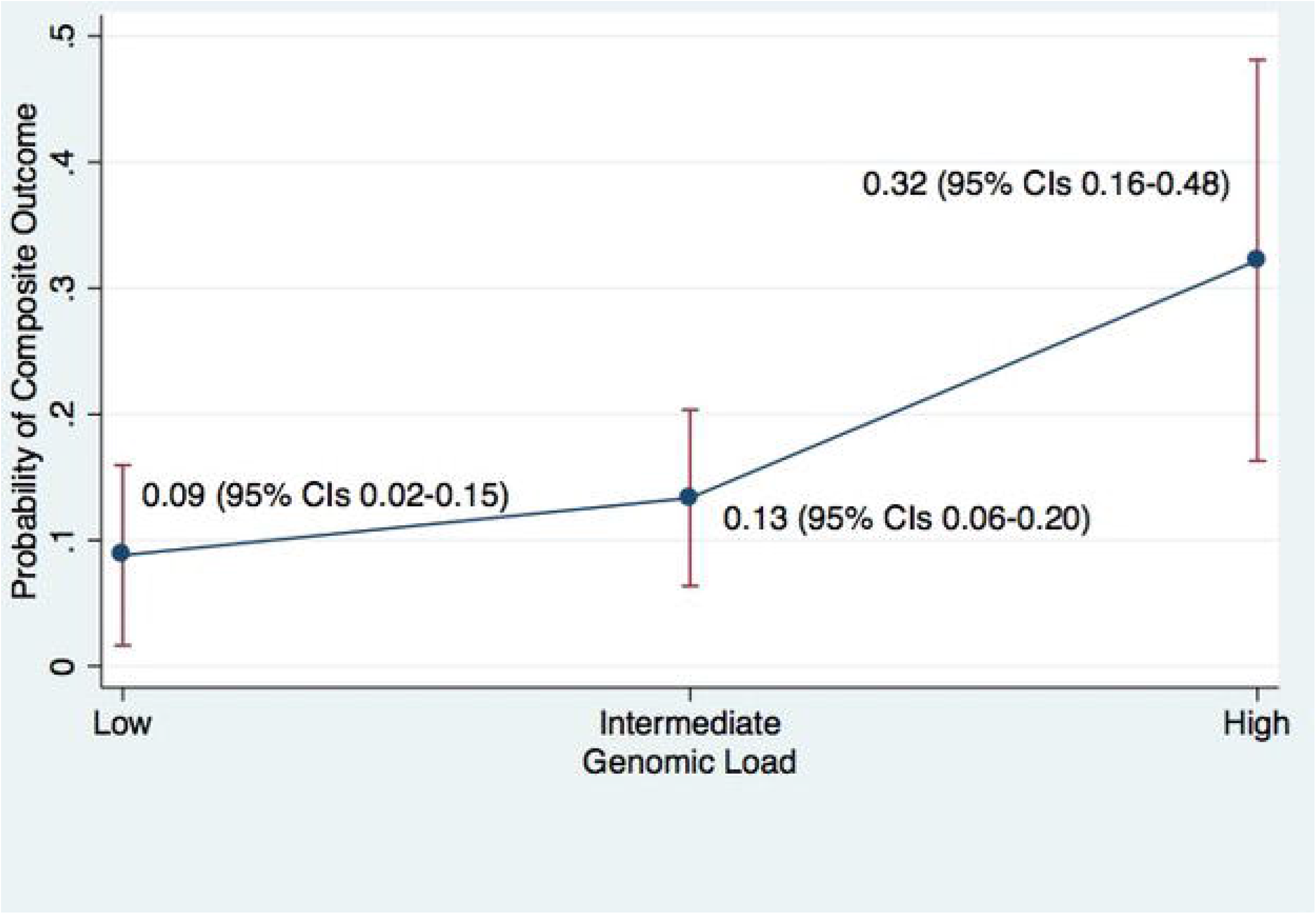
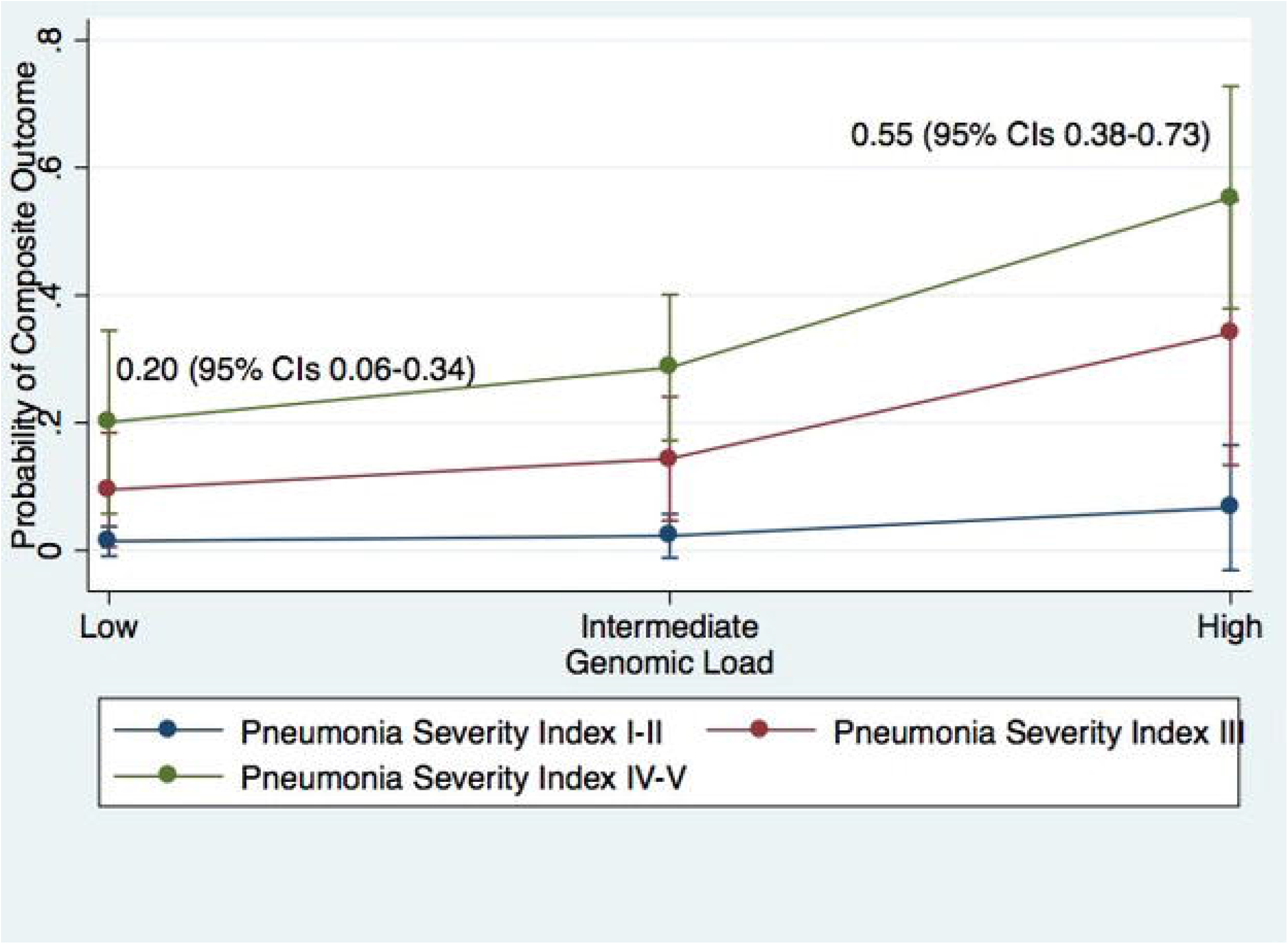
A. Prediction of Outcomes Based on Genomic Load. B. Prediction of Outcomes Based on Genomic Load and Pneumonia Severity Index.

## Discussion

We found that patients with a short duration of symptoms and high comorbidity index, as well as transplant recipients, were more likely to have a high SARS-CoV-2 genomic load at the time of hospital admission. The patients with high genomic load had a more severe clinical presentation and 3 times higher odds of dying or being intubated, independently of age, comorbidities, and severity of illness on presentation. Among patients with a severe clinical presentation at the time of hospital admission, patients with high genomic load were more than twice as likely to die or get intubated. This association remained significant on sensitivity analyses that included ARDS in the composite outcome and at varying Ct classification breakpoints.

Even though host factors have been shown to have an important role in COVID-19 outcomes (5–8), the contribution of viral factors in disease severity is less understood. The association of higher genomic load with poor clinical outcomes has been examined previously for other respiratory viruses (12). Previous studies on SARS-CoV-2 reported a similar viral load in respiratory specimens between asymptomatic and symptomatic patients (10, 20). Limited supportive data for the importance of SARS-CoV-2 viral load on disease severity have recently become available. Zheng *et al*. reported that among 96 patients with COVID-19, those with severe disease had significantly longer median duration of virus detection and higher viral loads in sputum samples compared to those with mild disease (21). Liu *et al*. reported that among 12 infected patients the viral load on throat swabs and bronchoalveolar lavage was associated with the development of ARDS (22). In another recently published study from China, among 48 admitted patients with COVID-19, detection of SARS-CoV-2 RNA in the serum was associated with higher serum levels of IL-6 and severe disease (23). The limited number of participants in these studies hasn’t allowed controlling for other factors that are known to influence outcomes, like age and comorbidities, while the various definitions for disease severity used limit generalizability of outcomes. To our knowledge, our study is the first to look into the association of genomic load in nasopharyngeal swabs collected on presentation per standard of care with robust clinical outcomes of death and intubation adjusting for a variety of possible confounding factors, such as patient demographics, comorbidities, and disease severity.

In our study, we examined the utility of genomic load from the upper respiratory tract in making inferences for the severity of pneumonia. We did this to evaluate if readily available Ct values of SARS-CoV-2 from nasopharyngeal samples at the time of initial patient evaluation for risk stratification could provide actionable results, in an era where appropriate triage is essential. While still unclear, it is plausible that lower respiratory samples may be more closely associated with clinical outcomes than nasopharyngeal samples (24). However, the difficulty in obtaining such samples makes it unlikely that this will be of significant value in daily clinical practice. Recent evidence suggests that there is active replication of SARS-CoV-2 in the upper respiratory tissues during the first five days after the onset of symptoms (25); a finding that correlates with our observation of higher genomic load in patients presenting within 7 days of symptom onset and substantiates our observation of significant association of nasopharyngeal genomic load with patient outcomes.

Another notable finding in our study was that patients with high genomic load were more likely to have a history of transplant with a trend seen among transplant patients for increased risk of intubation and death after adjustment for other comorbidities and demographic characteristics. This is the first study indicating transplantation as an independent factor for adverse outcomes and supports prior observations in cases series of a higher mortality rate among kidney transplant recipients compared to the reported mortality rate in the general population (26). There were few patients with other types of T cell immunodeficiency, such as those on chronic steroids or HIV, in our study population thus precluding any valuable conclusions.

Limitations of this study should be acknowledged and arise primarily from its retrospective design. Information was obtained from chart review, and as such incomplete reporting of patients’ characteristics and history of present illness is a consideration. However, both the primary outcome and the genomic load are objective measures that would not be influenced by incomplete reporting. Second, this study relies on Ct values obtained through a single assay, and the generalizability of the outcomes across different RT-PCR methods should be examined. Next, variation in the technique of obtaining the nasopharyngeal swab or collection of the specimen at different phases of the respiratory cycle could potentially cause fluctuation in the genomic load detected by the assay. These fluctuations would not be expected to result in systematic error and would have favored the null hypothesis. Finally, only the results of PCR-assay collected within 24 hours of admission were used in our analysis. Future studies should examine if a change in viral load during hospitalization is associated with a measurable effect in clinical status.

In summary, we showed that SARS-CoV-2 genomic load is an independent predictor of adverse outcomes in patients admitted to the hospital with COVID-19 pneumonia. Our findings suggest that antiviral therapy effective in reducing viral replication could potentially lead to improved outcomes. Importantly, we provide an independent predictor of adverse outcomes, that above and beyond age, comorbidities, and severity of illness on presentation, may be used to risk-stratify patients, in an era where appropriate triaging is of utmost importance. Finally, our findings call for a quantitative diagnostic assay to become available and future prospective studies to examine the effect of the use of viral load in re-admission rates and patient outcomes.

## Data Availability

Raw data were generated at NYU Langone Health, New York. Derived data supporting the findings of this study are available from the corresponding author on request.

## Acknowledgments

We thank all NYU Langone Health healthcare providers and laboratory technologists for their selfless devotion to patient care during the COVID-19 pandemic.

